# Temporal placental genome wide expression profiles reflect three phases of utero-placental blood flow during early to mid human gestation

**DOI:** 10.1101/2020.06.25.20139709

**Authors:** James Breen, Dale McAninch, Tanja Jankovic-Karasoulos, Dylan McCullough, Melanie D Smith, K Justinian Bogias, Qianhui Wan, Awais Choudhry, Nhi Hin, Stephen M Pederson, Tina Bianco-Miotto, Claire T Roberts

## Abstract

During early human placental development, extravillous cytotrophoblasts (EVT) invade the uterine vasculature to sequester a maternal blood supply. The impact of this on placental gene expression has not been established for normal pregnancy. Using RNA sequencing, we profiled placental chorionic villous tissues from 96 pregnancies at 6-23 weeks of gestation. We identified 1,048 genes that were differentially expressed between 6-10 weeks’ and 11-23 weeks’ of gestation. These are predominantly genes that are enriched in transcription factor signalling, inflammatory response and cell adhesion. Using a co-expression network and gene set enrichment analyses, we reveal three distinct phases of gene expression coincident with phases of maternal blood flow to the placenta that impact immune function and are likely driven by oxygen tension, potentially in a sex-specific manner. These data represent a comprehensive transcriptional profile of early placental development and point to significant environmental, genetic and regulatory triggers that drive gene expression.

## Main Text

### Introduction

Proper placental function is critical for the healthy development of the growing fetus *in utero*, and has a significant impact on long-term health of both the mother and child(*1*). Despite the critical role it plays during pregnancy, placental development is understudied, as are the molecular pathways controlling its function, mainly due to the difficulty in sampling placentas in ongoing pregnancies and the lack of well-established human placenta models(*2*). Placental key functions, however, are well established and include maternal-fetal exchange of nutrients, oxygen and wastes, and secretion of various peptides and steroid hormones that affect maternal adaptations to pregnancy and maintain maternal immune tolerance of the conceptus. Its diverse functions are attributed to its heterogeneous nature, with the proportion of individual cell types dynamically changing over time and with region. These include undifferentiated and differentiated cytotrophoblasts, syncytiotrophoblast with additional immune, endothelial and mesenchymal cells(*1*). All these cells work in concert with one another to support placental function which changes across gestation to accommodate different requirements in mother and fetus. Impaired placental differentiation and function can compromise pregnancy outcome. For example, inadequate placental extravillous cytotrophoblast (EVT) invasion, colonisation and transformation of the uterine spiral arterioles in early gestation can result in restricted placental blood flow and lead to pregnancy complications such as preeclampsia, intrauterine growth restriction, preterm labor, preterm premature rupture of membranes, late spontaneous abortion and placental abruption(*3*).

A critical period for placental development occurs at 10-12 weeks of gestation and is characterised by a distinct change in resource delivery. Leading to this critical period, extravillous trophoblasts (EVTs) migrate interstitially, invading and transforming the uterine spiral arterioles in early gestation to sequester a utero-placental blood supply from the mother into the intervillous space(*4*). This process occurs over time with initial invasion commencing in the days and weeks following implantation and the formation of the cytotrophoblastic shell. EVT plugging of decidual spiral arterioles is present as early as 6 weeks’ gestation with further invasion into the myometrial segments of these vessels occurring from 14 weeks’ gestation(*4, 5*). Paradoxically, the spiral arterioles are occluded by EVT plugs until about 10 weeks’ gestation, making the developing placenta (and fetus) gestate in a relatively hypoxic state up to this time. Doppler ultrasound blood flow studies show that maternal blood flow into the placental intervillous space is initiated at about 10 weeks’ gestation and increases progressively in vessels from the placental periphery towards the centre over the following weeks(*1*). Thus establishment of utero-placental blood flow takes weeks to manifest. Direct measurements using oxygen probes show that oxygen tension within the placenta begins to rise from <20mmHg at 8 weeks’ to ∼55mmHg at 13-14 weeks’ gestation(*6*).

As a whole, the placenta expresses a significant number of genes thought to be tissue-enriched (>4 fold difference compared to other tissues) across 19 human tissue profiles(*7*), with only brain, testes and white blood cells containing more uniquely expressed genes than placenta. Conservation of placental gene expression and function has been demonstrated in 14 eutherian mammalian species. Specifically, a core group of 115 genes is expressed at functional levels (fragments per kilobase per million mapped reads; FPKM >=10) relating to immune-modulation, cell-cell interaction, invasion and syncytialization, all essential for conserved placental function(*8*). In humans, relatively high levels of insulin-like growth factor (IGF) 2, pregnancy-associated plasma protein A (PAPP-A) and hypoxia-inducible factor (HIF)2a are all characteristic of placental chorionic villous tissue(*9*), the latter being the dominant transcription factor responding to the chronic non-pathological hypoxia which is present in the early gestation placenta.

Detailed transcriptional profiling of early gestation placenta would improve our understanding of normal regulation of placental development which is critical to identification of what goes wrong early in development in pregnancy complications. However, most studies of placental gene expression lack sufficient samples spanning early-mid gestation. To address this resource and knowledge gap, we sampled and sequenced 125 early gestation (6-23 weeks) placentas obtained from Australian women undergoing elective terminations of otherwise healthy pregnancies in Adelaide, South Australia. We quantified and compared differential gene expression in 6-10 weeks’ versus 11-23 weeks’ gestation, and identified co-expressed gene modules that dynamically change with each gestational week.

## Results

Overall, an initial set of 96 NIH samples were sequenced to an average of ∼35.8 million paired-end reads per sample, with sample PAC025 removed due to a lack of sequencing coverage, leaving a total of 95 samples available for gene expression profiling (Table S1). After initial filtering and clustering of gene expression profiles, a multi-dimensional scaling (MDS) plot (Fig. S1) displayed a clear gradient of gestational age across the second dimension, and clear separation between first (6-12 weeks’ gestation) and second trimester (13-23 weeks’ gestation) groups. Significant variability across the first dimension characterized the first trimester placenta, with a group of 11 first trimester samples clustering separately from all other profiles based on the first dimension. These profiles were shown to partly result from inadvertent failure to completely separate maternal decidua tissue from early gestation placenta(*10*). Given the variability within these samples, we chose to remove them from downstream analyses leaving a total of 84 samples. Genes on the sex chromosomes were removed from the dataset to reduce confounding by sex-differences in gene expression. From the reduced gene and sample set, 7,981 genes had zero read counts across all samples. After excluding genes with <2 counts per million (CPM) across 55 or more samples, we used expression data for a total of 15,648 genes for downstream analyses.

### Placental gene expression profiles change significantly from early to mid gestation

There is strong support for a gradient of change in placental oxygen tension across the first trimester as a result of maternal blood flow to the placenta, which starts to occur around 10-11 weeks’ gestation(*6*). Given our samples span across this early period in placental development, we aimed to define transcriptional profiles from otherwise normal pregnancies across early to mid-gestation to establish a molecular basis for placental functional changes across time.

When visualising normalised gene counts and samples using an PCA plot (Fig. 1a), we observed a gestational age gradient where samples were spread over the first dimension starting from early gestation (6 weeks’ PC1 ∼ −0.2) and ending with two samples at the other end of the gestational age spectrum (21/23 weeks’, PC1 ∼ 0.15). The top 50 most variable genes from PC1 (Fig. 1b) also split roughly across gestation, with seven variable genes showing large differences across this timepoint reflecting putative changes in oxygenation. These include the genes encoding glucose transporter 1 (SLC2A1), Kunitz-type serine proteinase inhibitor (TFPI2), epsilon globin 1 (HBE1), type I collagen 2 (COL1A2), antagonist of angiopoietin 1 and endothelial TEK tyrosine kinase (ANGPT1/TIE-2/TEK), chorionic somatomammotropin hormone 2 (CSH2) and pregnancy-associated plasma protein A (PAPPA). All seven genes are highly expressed in placenta and clustered together with a change from high gene expression in early gestational weeks to lower expression at later stages. Conversely, genes such as fibronectin 1 (FN1), growth differentiation factor 15 (GDF15) and type I collagen alpha 1 (COL1A1) also showed high placental expression in previous studies but showed an increase in gene expression across gestational weeks 6-23.

**Fig 1:**
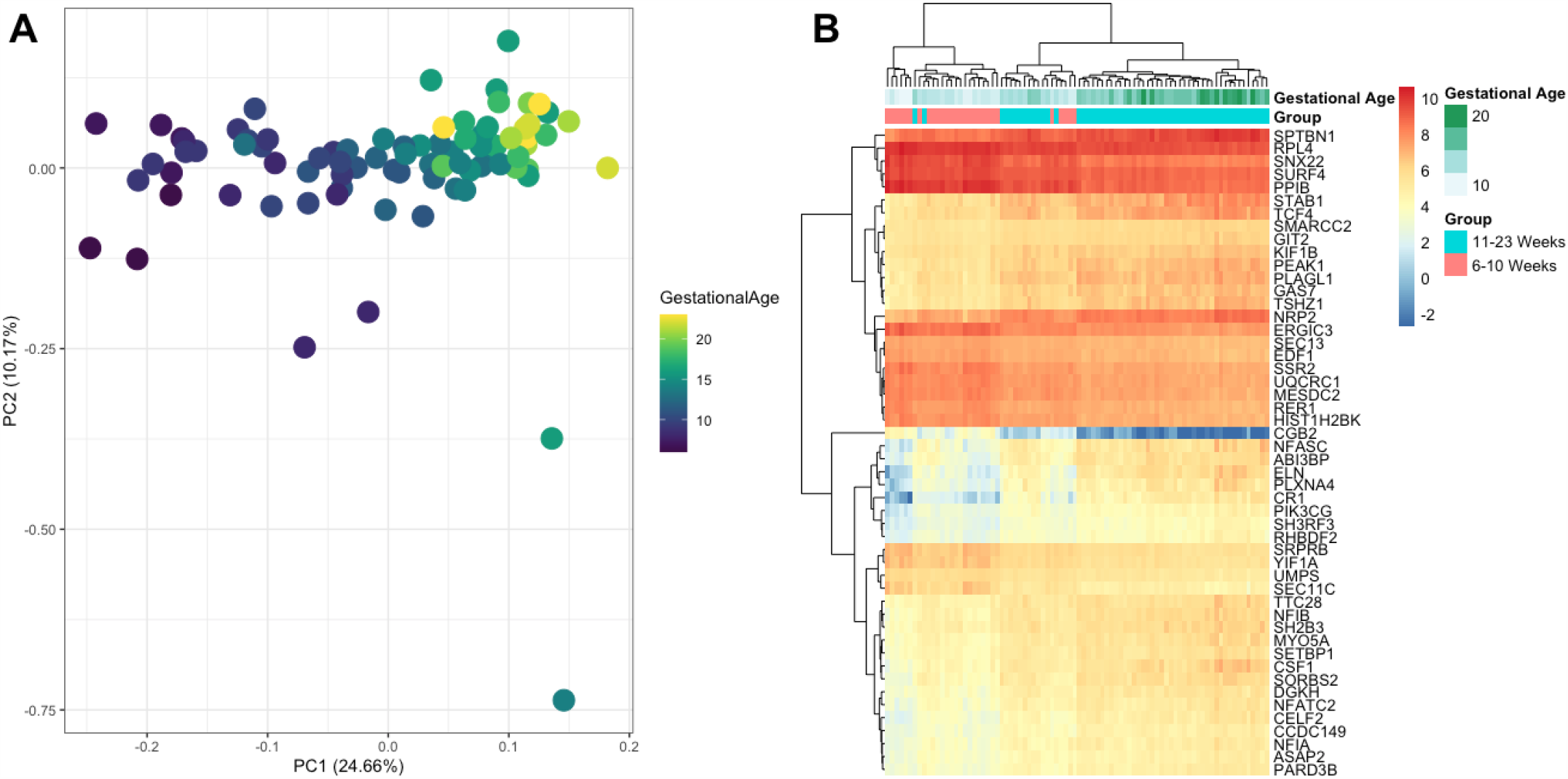
Unsupervised clustering of 84 placental RNA-seq samples across gestation from 6-23 weeks. (a) Principal Component Analysis (PCA) plot of all samples. (b) Heatmap of the top 50 genes from the log2 gene counts (CPM) that contribute to PC1. Annotations at the top of the heatmap indicate gestational age and groups before (6-10 weeks’) and after (11-23 weeks’) the putative initiation of oxygenated maternal blood flow into the intervillous space.

### Differentially expressed genes are enriched for vascular development early in pregnancy and immune response after 11 weeks’ gestation

After observing clear changes across the 10-11 weeks’ gestation threshold (Fig. 1a), we then conducted differential gene expression (DE) analyses over two gestational age groups; 6-10 weeks’ and 11-23 weeks’ gestation. After differential expression analyses using sample weights on log transformed filtered counts, we identified 8,544 genes that were differentially expressed according to gestation groups, with 4084 up-regulated and 4,460 down-regulated genes. Using additional log2 fold change filtering (logFC +/-1) and removing genes without adequate gene annotation we identified a final list of 1,048 genes that were significantly DE, 747 of which were up-regulated (i.e. higher expression after 10 weeks’ gestation) and 301 that were down-regulated (Fig. 2). The top 10 most differentially expressed genes across early to mid-gestation groups are shown in Table 1.

**Table 1:** Top 10 up and down-regulated differentially expressed genes between the 6-10 weeks’ and 11-23 weeks’ gestational groups. Genes are ordered in the table by fold change (logFC).

Upregulated (Higher in the 11-23 weeks' gestation group)
| Gene Name | EnsemblID | Gene Biotype | logFC | AveExpr | FDR |
| --- | --- | --- | --- | --- | --- |
| ALPP | ENSG00000163283 | Protein Coding | 4.31 | 6.57 | 4.11E-17 |
| GABRA4 | ENSG00000109158 | Protein Coding | 4.06 | -0.89 | 1.78E-08 |
| CD209 | ENSG00000090659 | Protein Coding | 3.90 | 0.82 | 2.88E-08 |
| MSC | ENSG00000178860 | Protein Coding | 3.59 | 2.67 | 1.34E-11 |
| CR1 | ENSG00000203710 | Protein Coding | 3.59 | 3.37 | 3.66E-15 |
| FGF10 | ENSG00000070193 | Protein Coding | 3.50 | 0.86 | 4.09E-06 |
| LINC00523 | ENSG00000196273 | lincRNA | 3.42 | -0.38 | 1.69E-10 |
| ELN | ENSG00000049540 | Protein Coding | 3.23 | 4.25 | 5.78E-16 |
| WNT10A | ENSG00000135925 | Protein Coding | 3.21 | 1.74 | 3.95E-16 |
| TMEM176B | ENSG00000106565 | Protein Coding | 3.20 | 3.89 | 2.32E-10 |

Downregulated (Higher in 6-10 weeks' gestation group)
| Gene Name | EnsemblID | Gene Biotype | logFC | AveExpr | FDR |
| --- | --- | --- | --- | --- | --- |
| HBE1 | ENSG00000213931 | Protein Coding | -6.48 | 4.62 | 2.30E-25 |
| HBZ | ENSG00000130656 | Protein Coding | -6.04 | 1.09 | 1.11E-16 |
| MMP12 | ENSG00000110347 | Processed Transcript | -5.44 | 2.90 | 1.97E-15 |
| ANGPT4 | ENSG00000101280 | Protein Coding | -5.21 | 0.11 | 1.80E-16 |
| CGB2 | ENSG00000104818 | Protein Coding | -4.97 | -0.34 | 1.73E-18 |
| C7orf71 | ENSG00000222004 | Protein Coding | -4.89 | 0.97 | 4.39E-17 |
| GPR32 | ENSG00000142511 | Protein Coding | -4.66 | 2.87 | 4.05E-15 |
| SLC6A19 | ENSG00000174358 | Protein Coding | -4.54 | -0.68 | 2.94E-16 |
| CGB7 | ENSG00000196337 | Protein Coding | -4.52 | 3.89 | 8.50E-17 |
| PAEP | ENSG00000122133 | Protein Coding | -4.33 | 3.92 | 3.35E-11 |

**Fig 2:**
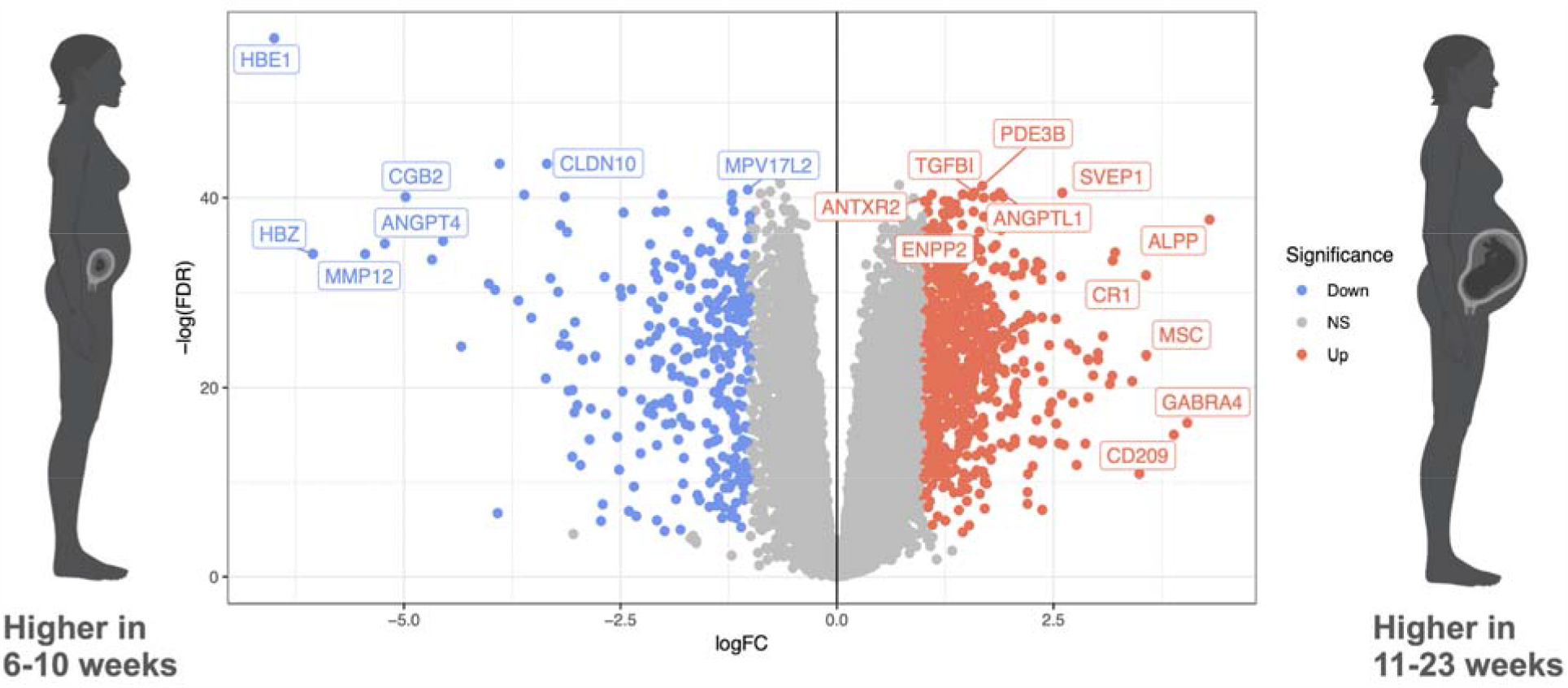
Differentially expressed genes (FDR<0.05 & logFC +/-1) transecting the putative time at which maternal blood flow to the developing placenta is initiated. Volcano plot showing the level of change (log transformed normalised counts) and differential expression of genes observed between 6-10 weeks’ gestation and 11-23 weeks’ gestation.

There were 446 more down-regulated DE genes than up-regulated when comparing groups. Down-regulated genes after 10 weeks’ gestation displayed a greater variance in fold change compared to up-regulated genes, with five genes (HBE1, HBZ, MMP12, ANGPT4) logFC < -5, with hemoglobin subunit epsilon 1 gene (HBE1) being the most highly down-regulated (-6.5) as well as having the greatest significant change (FDR = 2.16e^-25^) across all genes. Chromosome 19 chorionic gonadotropin subunit beta genes (CGB, CGB2, CGB5, CGB7 and CGB8), that are expressed by trophoblasts in early gestation, were all highly down-regulated (<-2.98 logFC) and highly expressed, with the exception being CGB2 that had low expression but was the most significantly different between 6-10 weeks’ and 11-23 weeks’ gestation. Conversely, only two up-regulated genes had a greater than +4 logFC difference, with placental alkaline phosphatase gene (ALPP) being the highest up-regulated gene (+4.3 logFC). Taken together, significant DE genes were all found to have high expression in placental tissue in previous studies and public reference expression sets(*11*). The majority of highly significantly DE genes have unique expression in placenta or limited expression in other sampled tissues. One exception, however, was the differentially expressed gene with the lowest significance (FDR 2.30e^-25^) human haemoglobin epsilon globin gene 1 (HBE1), which has expression in a range of tissues.

Gene ontology (GO) and functional gene set enrichment tests of DE genes between 6-10 weeks’ and 11-23 weeks’ gestation groups showed significant enrichment (FDR < 1e^-20^) in GO Biological Processes such as inflammatory response, immune system process, response/regulation of response to external stimulus and cell activation. Additionally, DE genes were enriched (FDR<1e^-20^) in plasma membrane and extracellular regions of the GO Cellular Components, and GO Molecular Functions were also enriched (FDR<1e^-09^) for genes relating to signal receptor, glycosaminoglycan and sulfur compound-binding (Table S2). Using KEGG pathway and Hallmark gene sets from the Molecular Signature Database (MSigDB; Table S3), DE genes were mostly enriched in pathways and gene sets relating to immune-response. This enrichment in each analysis was mainly driven by up-regulated genes, which were more likely to be enriched in immune response pathways.

### Gene co-expression analysis defines three distinct phases of early to mid gestation gene expression

While changes in immune regulation response were observed in the differential expression analyses, comparison between two gestational age groups (6-10 weeks’ & 11-23 weeks’) limits the ability to identify fine-scale transcriptional changes across the full 17 weeks’ time span that was sampled in this study. Therefore, we chose to establish a co-expression network of all available chorionic villous samples across early to mid gestation and correlate co-expressed genes to each gestational week between 6-23 weeks’ gestation to determine whether gene expression changes are continuous across time and whether the estimated initiation of maternal blood flow to the placenta can be discerned from RNA-seq data.

To enable greater resolution across first trimester, we included 26 additional (all 6-12 weeks’ gestation), high-coverage (average 45.2 million reads per sample) chorionic villous samples sequenced in a previous study (Mayne et al. *Unpublished*) in the co-expression analysis. All 110 samples (49 Female/61 Male) spanned 6-23 weeks’ gestation as for the differential expression analysis above (Table S1). After filtering for low expression (same methodology as previous dataset), genes with low variability and adjusting for batch effects, we used a core set of 10,197 genes for co-expression analyses. Samples showed similar MDS clustering to the previous dataset (Fig. S2), with samples clustering over a gradient of low-high gestational age, indicating that expression profiles would be comparable in co-expression analyses.

After network construction, we identified 17 modules of co-expressed genes containing 9,226 genes, which were then reduced to 14 after merging modules on a 0.15 cut height from the clustering dendrogram (Fig. S3). While most modules contain very specific co-expression gene clustering, the largest module (grey) contains unassigned genes from across the network and therefore was disregarded for downstream analyses. The remaining 13 modules (Fig. 3a) ranged in size from the largest composed of 1,060 genes (greenyellow), to 114 genes in the smallest (lightcyan), with the majority of modules containing an average of 350 genes (Table. 2).

**Fig 3:**
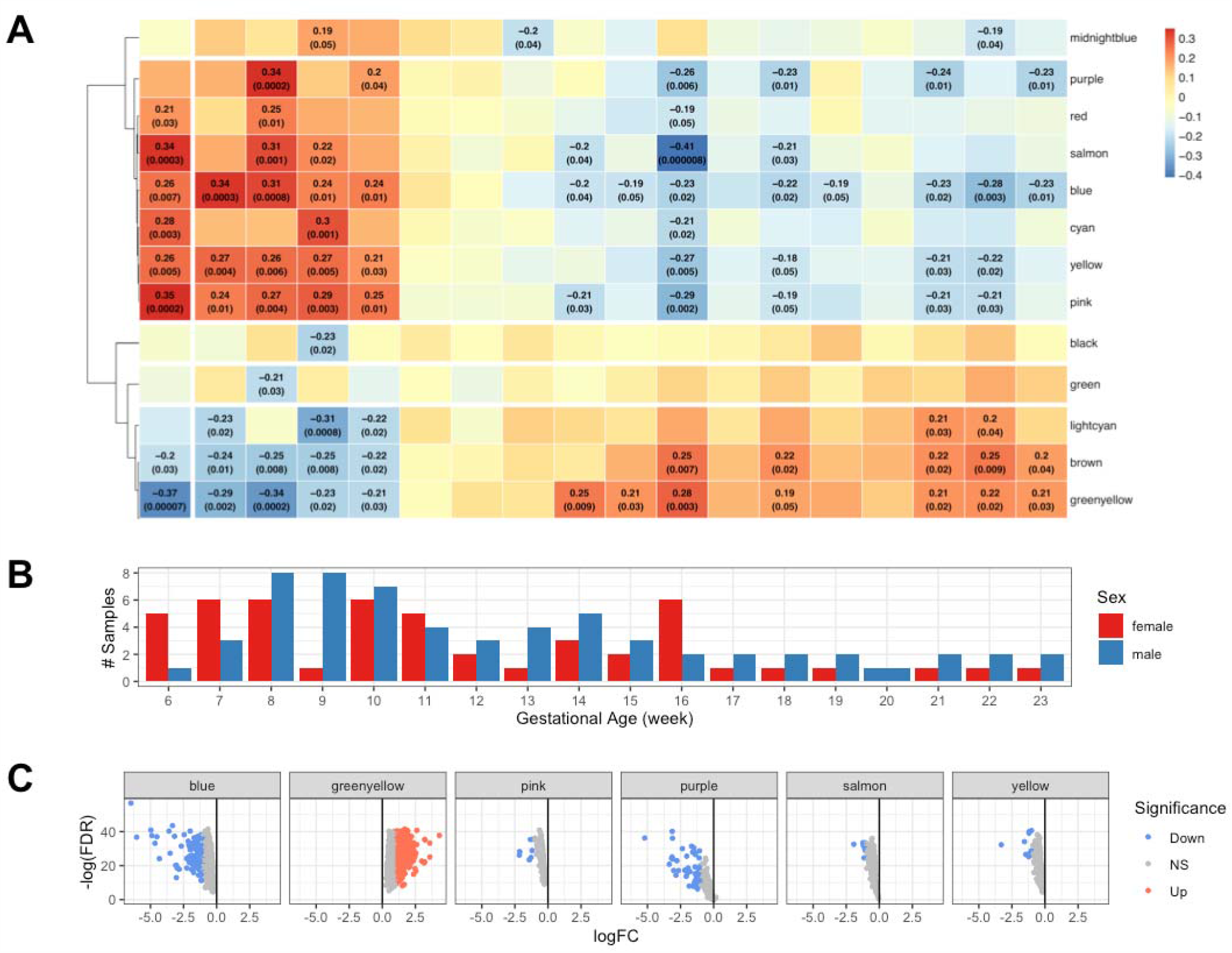
Whole Genome Co-expression Network Analysis (WGCNA) based on 110 chorionic villous samples identified 13 co-expressed gene modules (a) which were then correlated with 6-23 gestational weeks based on the sample distributions (with both male and female samples) displayed underneath the dendrogram (b). Module correlation to each gestational age (positive in red and negative in blue) is shown with only module/gestational week pairs being displayed if the correlation was equal to or lower than p-value 0.05. Six modules contained differentially expressed genes between 6-10 versus 11-23 weeks’ gestation (c).

Based on gestational week, correlation clustering patterns (Fig. 3ab), we identified a clear split in the gene modules correlated with early (6-10 weeks’) and later (14-23 weeks’) stages of placental development. Early weeks show positive correlation (correlation coefficient > 0) to 8 modules (midnightblue, purple, red, salmon, blue, cyan, yellow and pink). Genes found in these modules were shown to have significant (p-value ≤ 0.05) positive correlation, with module blue displaying significant positive correlation with each of the 5 weeks in that period (6-10 weeks’ gestation). Conversely, 5 modules (black, green, lightcyan, brown and greenyellow) have a distinctive negative correlation (correlation coefficient <0) with early gestational weeks. Each of these modules contained at least one week with significant negative correlation during 6-10 weeks’, with brown and greenyellow containing significant correlation in each of these weeks, although greenyellow had the greatest level of significance and negative correlation over this time. Interestingly, when looking at modules correlated with 14 and 23 weeks, the relationship is reversed, with highly positively correlated modules becoming negatively correlated with later gestational age and vice versa. While high/low correlation is observed in two different groups of modules at 6-10 and 14+ gestational weeks, very low correlation (-0.13 to 0.12) is observed in all 13 modules across 11-13 weeks’ gestation, with only one module midnightblue being negatively correlated (-0.2) with the week at a significant level. Thus 11-13 weeks’ gestation seems to be a transitionary period for placental transcription.

Given the co-expression modules positively and negatively correlate with opposite time-periods, before and after our differential expression time cut-off (10/11 weeks’), we then intersected DE genes with module membership information to identify the gene and level of expression that was differentially expressed (Table 2). Only 6 of the 13 modules contained DE genes (greenyellow, pink, purple, salmon, yellow and blue) shown in Fig. 3c, with greenyellow containing only genes that are upregulated between 6-10 and 11-23 weeks’ gestation groups. Greenyellow module contains genes that are positively correlated with later weeks’ gestation (14-23 weeks’), suggesting that genes found within this module are correlated with the immune system signature identified in the gene set enrichment analyses above. The other 5 modules all contained DE genes that were down-regulated.

**Table 2:** Numbers of total genes and differentially expressed (DE) genes found in each WGCNA module

| Module Name | Total Genes | DE Genes<br>(6-10 weeks' vs<br>11-23 weeks') |
| --- | --- | --- |
| black | 281 | - |
| blue | 436 | 106 |
| brown | 467 | - |
| cyan | 369 | - |
| green | 362 | - |
| greenyellow | 1060 | 526 |
| lightcyan | 114 | - |
| midnightblue | 118 | - |
| pink | 217 | 7 |
| purple | 199 | 52 |
| red | 289 | - |
| salmon | 280 | 9 |
| yellow | 360 | 11 |

We also constructed a network (Fig. 4) using co-expression connectivity to determine whether specific modules had additional connection to others. Nodes (genes) with less than 4 edges (co-expression level) to other nodes were removed leaving a network containing nodes primarily from *cyan, black, blue, purple, salmon, yellow* and *greenyellow* modules. The topology of the network is defined by a large cluster of *greenyellow* genes that are extremely well connected to genes within the same module, found in the top left of Fig. 4a. As shown before, these genes displayed significant enrichment in up-regulated immune response genes and also showed enrichment for GO categories such as cell signalling activation. Below and to the right of this module cluster is a group of 6 connected modules (*salmon, lightcyan, pink, purple, yellow* and *blue*), most of which are associated with early gestational age. Down-regulated DE genes were enriched in terms relating to RNA-binding, protein translation and processing, ribosomal synthesis, circulatory system development and mitochondrial activity (Table S4).

**Fig 4:**
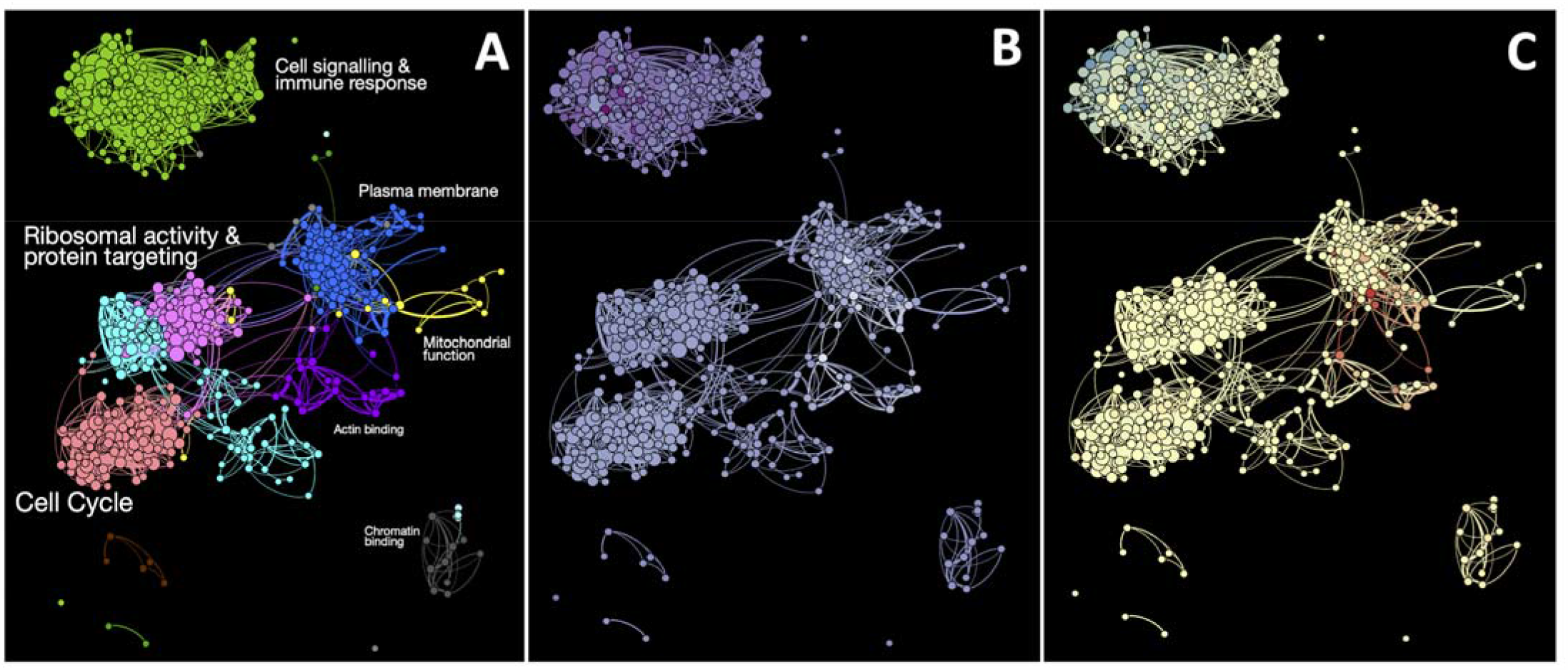
Gene networks identified through connectivity within each WGCNA module. Gene modules (a) were displayed along with enriched GO terms (font size scaled by adjusted p-value e.g. large text indicates higher significance). Nodes (b) were then coloured for fold change in differential expression analyses between 6-10 weeks’ and 11-23 weeks’ gestation, white being early in gestation and purple being later in gestation, and sex (c) where red is female-biased and blue is male-biased.

Genes from the *greenyellow* gene module had higher total connectivity (connectedness of all genes within the module) than all other modules (Fig. S4), with the more highly connected genes within the module being much more likely to be enriched in the immune response function. After identifying network connectivity associated with each gene module (Fig. 4a), we then overlayed differential expression results from the DE analysis (Fig. 4b) and autosomal sex-biased gene expression changes (Fig. 4c) to establish whether initiation and establishment of maternal blood flow to the placenta affected the connections between eigengenes. G*reenyellow* module immune system associated module genes displayed greater changes from later time periods (deeper purple colour of each gene), compared to genes found in other modules (Fig. 4b). Interestingly, genes showing autosomal gene expression bias towards one sex or the other were located within specific niches of the network that also had greater fold change in the DE analysis between 6-10 weeks’ and 11-23 weeks’ gestation (Fig. 4c).

Genes with lower connectivity were enriched with genes associated with vascular development, potentially due to EVT invasion and increasing chorionic villous vascularization after 11 weeks’ gestation. In order to invade, EVTs must undergo an epithelial to mesenchymal transition (EMT) process. EMT has been shown to be important in the formation of EVTs and ultimately the establishment of an adequate maternal blood supply to the placenta. Vasculogenesis in the placental villi would also be expected to be accompanied by a mesenchymal gene signature. To identify the relative state in which samples were found in this process, we utilised single-sample gene enrichment scoring(*12*) with established cell-line and tumour-derived EMT gene signatures(*13*). Using both normal and cancer-related cell-line gene signatures (Fig. 5a) we note an increase of expression from a characteristic epithelial signature (ES) to a mesenchymal signature (MES) from 6 to 23 weeks’ gestation. In both signatures, variance of expression (dispersion) decreases for ES in early gestation samples and MES in later samples, suggesting that a transition between the two cell states occurs during this phase in placental development where trophoblast invasion and placental fetal vascularization would dramatically alter gene expression, as previously shown in Fig. 3 and 4. Many genes within ES and MES sets were also found in adaptive and innate immune gene sets (Fig. 5b) which also overlap with *greenyellow* genes, indicating a shared pathway connection between immune function and EMT. Interestingly, tumour cell line derived genes showed a clearer clustering pattern from high to low dispersion in ES and MES signatures compared to normal cell-line dataset, indicating that tumour gene signatures are potentially more reflective of placental development, with the caveat being the variability of sampled genes in each case.

**Fig 5:**
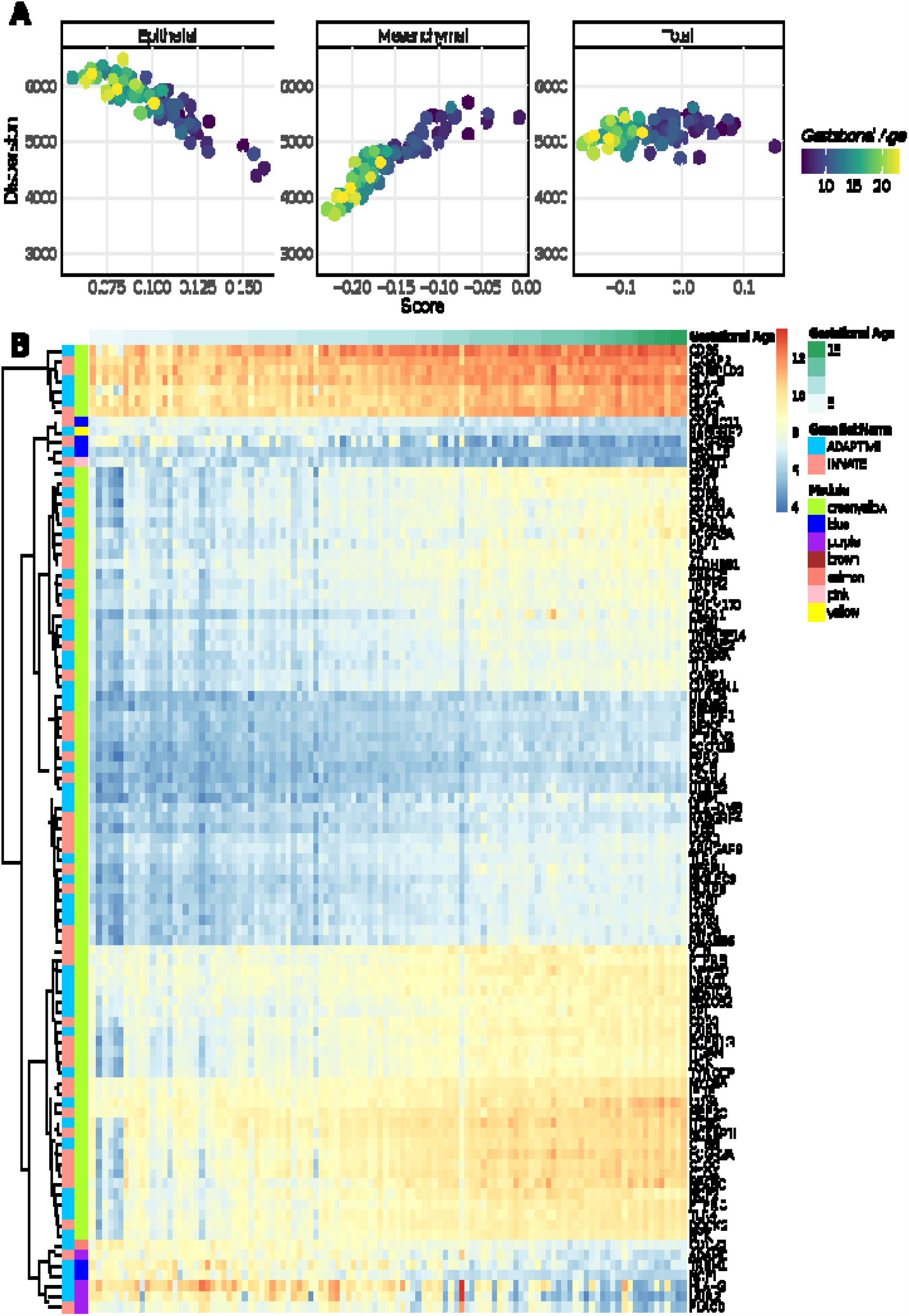
Enrichment of Epithelial-Mesenchymal Transition (EMT) and Adaptive/Innate Immune response genes across all samples. (a) Tumour-derived gene signatures were used across all samples using single-sample gene set enrichment. (b) Variable genes (>0.6 standard deviation in expression) from the Reactome Adaptive/Innate Immune Response genes from MSigDB gene sets were also displayed in a heatmap alongside module membership and sample metadata.

When investigating global gene expression across both gestational age and sex comparisons further, we find that 29 genes had a gestational age change greater/less than 1 logFC and sex-biased with logFC greater/less than 0.8. Interestingly, all genes are found to be biased in the same direction, with placenta from all female-bearing pregnancies showing down-regulation (i.e. expression in 6-10 weeks’ greater than 11-23 weeks’ gestation), and from all male-bearing pregnancies showing up-regulation (Fig. 6a and Table 3). Examination of co-expressed modules confirmed these results, as *greenyellow* genes had positive correlation with placenta from male-bearing pregnancies, with *blue, pink, purple* and *red* modules having the opposite effect (Fig. 6b).

**Table 3:**
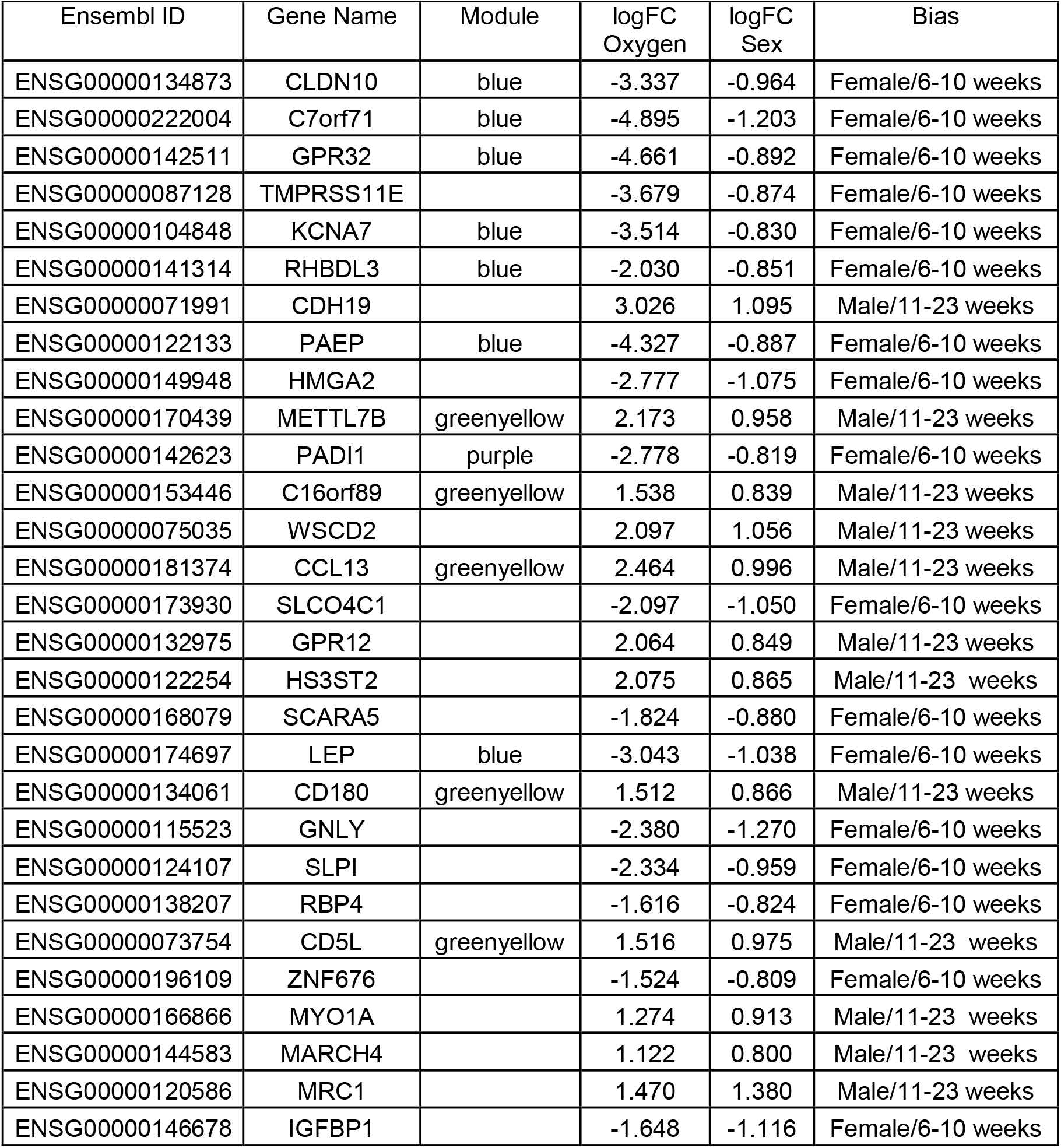
List of genes with higher fold change in both 6-10 weeks’ versus 11-23 weeks’ gestation and sex-biased group comparisons. All 29 genes with absolute logFC > 1 in the gestational age and absolute logFC > 0.8 in sex comparisons are shown with module information along with the bias directions (Male/Female, 6-10/11-23 weeks’ gestation).

| Ensembl ID | Gene Name | Module | logFC<br>Oxygen | logFC<br>Sex | Bias |
| --- | --- | --- | --- | --- | --- |
| ENSG00000134873 | CLDN10 | blue | -3.337 | -0.964 | Female/6-10 weeks |
| ENSG00000222004 | C7orf71 | blue | -4.895 | -1.203 | Female/6-10 weeks |
| ENSG00000142511 | GPR32 | blue | -4.661 | -0.892 | Female/6-10 weeks |
| ENSG00000087128 | TMPRSS11E |  | -3.679 | -0.874 | Female/6-10 weeks |
| ENSG00000104848 | KCNA7 | blue | -3.514 | -0.830 | Female/6-10 weeks |
| ENSG00000141314 | RHBDL3 | blue | -2.030 | -0.851 | Female/6-10 weeks |
| ENSG00000071991 | CDH19 |  | 3.026 | 1.095 | Male/11-23 weeks |
| ENSG00000122133 | PAEP | blue | -4.327 | -0.887 | Female/6-10 weeks |
| ENSG00000149948 | HMGA2 |  | -2.777 | -1.075 | Female/6-10 weeks |
| ENSG00000170439 | METTL7B | greenyellow | 2.173 | 0.958 | Male/11-23 weeks |
| ENSG00000142623 | PADI1 | purple | -2.778 | -0.819 | Female/6-10 weeks |
| ENSG00000153446 | C16orf89 | greenyellow | 1.538 | 0.839 | Male/11-23 weeks |
| ENSG00000075035 | WSCD2 |  | 2.097 | 1.056 | Male/11-23 weeks |
| ENSG00000181374 | CCL13 | greenyellow | 2.464 | 0.996 | Male/11-23 weeks |
| ENSG00000173930 | SLCO4C1 |  | -2.097 | -1.050 | Female/6-10 weeks |
| ENSG00000132975 | GPR12 |  | 2.064 | 0.849 | Male/11-23 weeks |
| ENSG00000122254 | HS3ST2 |  | 2.075 | 0.865 | Male/11-23 weeks |
| ENSG00000168079 | SCARA5 |  | -1.824 | -0.880 | Female/6-10 weeks |
| ENSG00000174697 | LEP | blue | -3.043 | -1.038 | Female/6-10 weeks |
| ENSG00000134061 | CD180 | greenyellow | 1.512 | 0.866 | Male/11-23 weeks |
| ENSG00000115523 | GNLY |  | -2.380 | -1.270 | Female/6-10 weeks |
| ENSG00000124107 | SLPI |  | -2.334 | -0.959 | Female/6-10 weeks |
| ENSG00000138207 | RBP4 |  | -1.616 | -0.824 | Female/6-10 weeks |
| ENSG00000073754 | CD5L | greenyellow | 1.516 | 0.975 | Male/11-23 weeks |
| ENSG00000196109 | ZNF676 |  | -1.524 | -0.809 | Female/6-10 weeks |
| ENSG00000166866 | MYO1A |  | 1.274 | 0.913 | Male/11-23 weeks |
| ENSG00000144583 | MARCH4 |  | 1.122 | 0.800 | Male/11-23 weeks |
| ENSG00000120586 | MRC1 |  | 1.470 | 1.380 | Male/11-23 weeks |
| ENSG00000146678 | IGFBP1 |  | -1.648 | -1.116 | Female/6-10 weeks |

**Fig 6:**
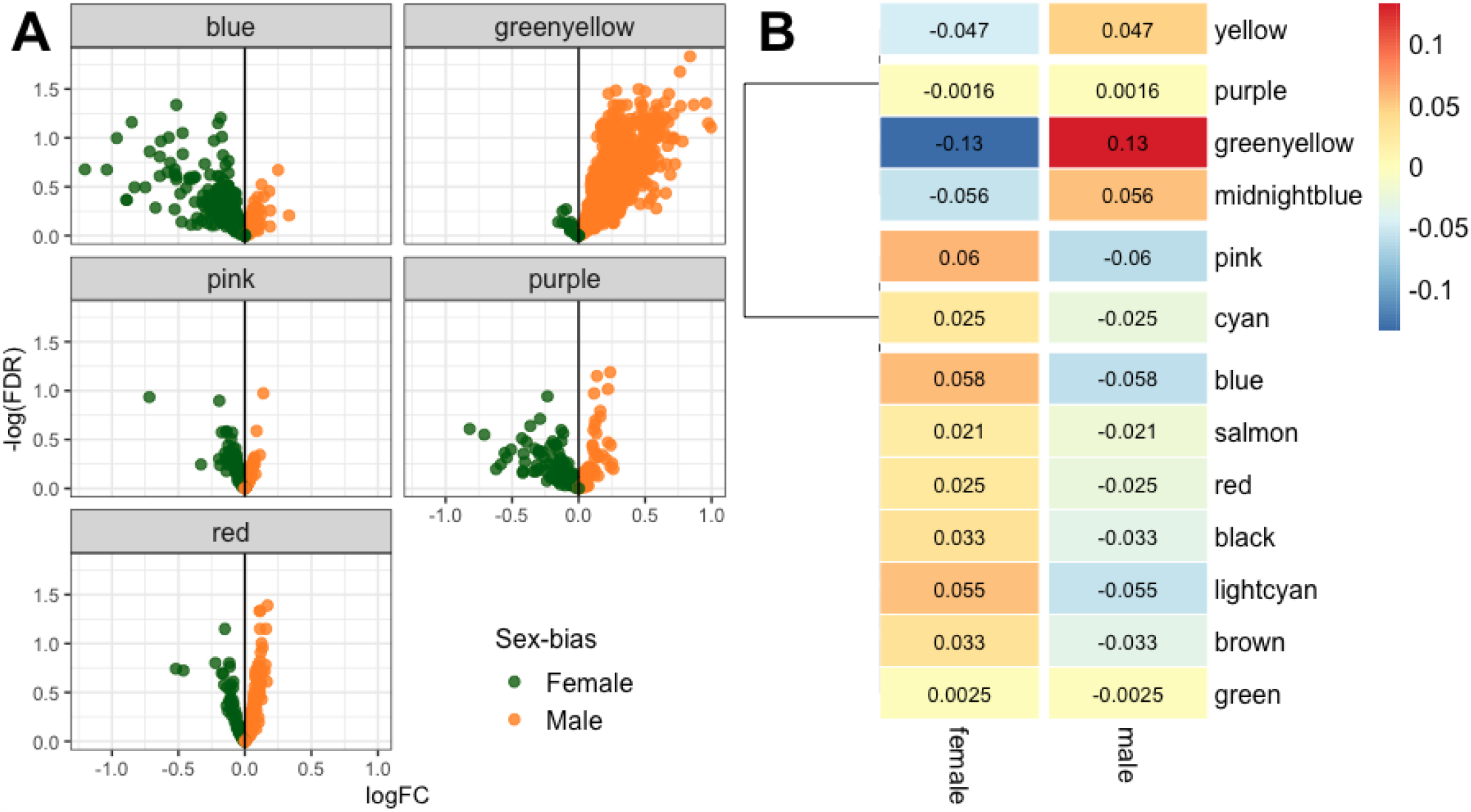
Sex-biased gene expression identified in RNA-seq data. Volcano plots of sex-biased expressed genes in each WGCNA module that contained at least one gene greater or less than - 0.5/0.5 displayed sex-biased expression patterns (a), with greenyellow module genes having the greatest correlation with male-bearing pregnancies (b).

## Discussion

Genome wide expression profiling provides an invaluable snapshot to define the range of biological functions that occur within a tissue at a particular point in time. It can be used to understand molecular mechanisms in organ function and identify potential diagnostic and therapeutic targets in disease states. Characterising functional gene expression in the placenta is arguably more challenging than for other tissues due to its developmental dynamic changes in structure and function across gestation. Early gestation placenta is largely understudied due to perceived ethical issues surrounding the use of pregnancy termination samples in some jurisdictions. The ideal would be to undertake longitudinal serial placental sampling across gestation in ongoing pregnancies but the inordinate risk associated with this is prohibitive. Unfortunately, a good non-primate animal model of human placenta is lacking(*2*).

Recent studies have investigated placental gene expression in early gestation(*14*–*16*), including at a single cell level(*17*–*19*), indicating that gene expression profiles at the maternal-fetal interface reflect a diverse array of cell populations from maternal and fetal lineages.

To our knowledge, our study provides the highest resolution of the placental transcriptome across the first and second trimester conducted to date. RNA sequencing of large numbers of early to mid gestation placenta has allowed us to detail dynamic changes in the transcriptome in presumably normal placental development and highlight potential sex-biased expression of genes associated with maternal-fetal circulatory changes.

The pattern of gene expression highlighted herein demarcates three distinct temporal phases to mid gestation; early gestation (6-10 weeks’), a transition phase (11-13 weeks’) and early second trimester to mid gestation (14-23 weeks). Initially, unsupervised clustering of filtered gene expression profiles (Fig. 1a) showed a clear gradient of expression from the earliest to latest gestations studied, indicating that temporal status drives gene expression variation. This change is similar to that shown previously for first and second trimester placental DNA methylation(*20*).

The first phase of placental gene expression, from 6-10 weeks’ gestation (Fig. 3) that is characterised by high expression of genes containing attributes of protein processing, ribosomal synthesis and cell-cycle functions (Fig. 4a). This phase equates with the period of essentially absent utero-placental blood flow due to occlusion of the spiral arterioles by invading EVTs. However, during this time vasculogenesis stimulated by vascular endothelial growth factor (VEGFA) begins to occur(*21*) consistent with gene enrichment of plasma membrane GO terms and angiogenesis genes (e.g. ANGPT4) seen in our study from 6-10 weeks’ gestation. Co-expressed gene modules with positive correlation with gestational weeks 6-10 were also enriched in membrane and cell adhesion gene ontologies. Cell populations defined in single cell RNA-seq profiles during this stage indicate that expression of these genes is partly driven by fusion of villous cytotrophoblasts to syncytiotrophoblast or by differentiation of extravillous trophoblasts(*19, 20, 22, 23*). Although there is a dominance of epithelial gene signatures, epithelial-mesenchymal transition (EMT) gene signature analysis shows some mesenchymal gene expression which may associate with EMT in distal cytotrophoblast shell differentiation to EVT but also differentiation of chorionic villus mesenchyme with early capillary formation.

From 11 weeks’ gestation, the profile of gene expression dynamically changes to an immune response state, first going through a transition phase between 11 and 13 weeks’ gestation, and then undergoing a complete gene signature shift from 14 weeks’ until the end of our sampling period at 23 weeks’ gestation (Fig. 3). The late first trimester transition phase occurs at the same time that maternal blood flow to the placental intervillous space initiates and becomes established. This transitional phase of gene expression revealed by WGCNA, is characterised by an up-regulation of genes containing attributes of the immune response. This is apparent in both the up-regulated DE genes from 11 weeks’ gestation and in the large numbers of co-expressed genes found in the greenyellow co-expressed gene module. Up-regulation of immune response genes has previously been linked with second trimester placental function(*19, 20, 22, 23*). scRNA-seq research suggests that placental adaptive and innate immune responses are essential to prevent the maternal immune system attacking the antigenically ‘foreign’ conceptus(*19*). Given the current scRNA-seq resources available from early pregnancy and term placental tissue (*17*–*19, 23*), there is significant potential to link single cell assessment of placental gene expression to the identified changes in phenotype identified in our study.

The identification of sex-biased expression of autosomal genes linked to differential placental gene expression between 6-10 weeks’ and 11-23 weeks’ gestation is interesting. Here we show down-regulation of gene expression in female-bearing pregnancies across the 10-11 weeks’ threshold with up-regulation, particularly of immune response genes, in male-bearing pregnancies (Fig. 6). We have previously shown sex-biased expression in term placenta from uncomplicated pregnancies(*24*).

Sexual dimorphism in fetal growth and pregnancy complications has long been established(*25*– *27*), with male fetuses having a greater risk of being born extremely preterm and to a pregnancy complicated by gestational diabetes, while female-bearing pregnancies have greater risk for early onset hypertensive disorders of pregnancy(*25*–*27*). Hormone differences may account for much of the expression differences seen in female placenta samples in our study. However, the high connectivity within immune system genes observed to be male-biased point to the potential that dysregulation of immune tolerance may account for fetal sex-bias in pregnancy complications associated with inflammation that arise later in gestation. Variations in fetal sex-biased placental gene expression observed in this study are likely to be an under-estimation of the full complement of sexually dimorphic placental gene expression, given that we excluded data from genes on both sex chromosomes from our analyses.

Our data show that genome wide expression profiles in placenta from 6-23 weeks’ gestation largely reflect an early phase of relative hypoxia due to little to no utero-placental blood flow; followed by a transition phase that reflects initiation of and increasing maternal blood flow to the placenta over a period of about four weeks (11-13 weeks’); with final establishment of the utero-placental circulation from 14-23 weeks’ gestation. Hence, these three phases also likely reflect oxygen tension within the placenta which is known to be a powerful regulator of gene transcription. The datasets generated in this study offer an important reference set of gene expression profiles for future studies, especially when coupled with profiles from pregnancies with underlying complications.

## Materials & Methods

### Experimental design and ethics statement

To establish gene expression profiles across early gestation, first and second trimester placental tissue (6-23 weeks’ gestation) were collected from women undergoing elective terminations of otherwise healthy pregnancy at the Women’s and Children’s Hospital North Adelaide and the Pregnancy Advisory Centre, Woodville, Adelaide. Written, informed consent was obtained from all patients prior to collection of placental tissue. Collection of first and second trimester placental tissue was approved by the Women’s and Children’s Health Network Human Research Ethics Committee (REC2249/2/13), University of Adelaide Human Research Ethics Committee (H-137-2006), Queen Elizabeth Hospital and Lyell McEwin Hospital Human Research Ethics Committee (REC 1712/5/2008 and HREC/12/TQEHLMH/16).

### Sample Information

To characterise placental gene expression across early to mid gestation, we initially sampled and sequenced 96 chorionic villous placenta samples spanning gestational ages 6-23 weeks collected from women having non-medical elective terminations from otherwise normal pregnancies (“NIH” dataset). Samples were genotyped for sex, with 44 and 51 female and male-bearing pregnancies respectively, and gestational weeks from early to mid gestation. An additional 29 early gestation placentas from pregnancy terminations from 6-11 weeks’ gestation (14 female and 15 male-bearing pregnancies) collected from the Women’s and Children’s Hospital in North Adelaide were also used later for co-expression analyses (“ADEL” Dataset), with the same wet laboratory work (RNA extracted and library preparation) and sequence analysis methods carried out on each, albeit separated by multiple years.

### RNA extraction, library preparation and sequencing

Placental villous tissue was collected from women who were undergoing elective pregnancy terminations (6-23 weeks’ gestation) immersed in RNAlater solution (Invitrogen) within minutes of termination and placed at -4°C for 24 h prior to being stored at -80°C. RNA was extracted from each placental villous sample using the Qiagen RNeasy kit following the manufacturer’s protocols. Sequencing libraries were prepared using Illumina TruSeq Stranded Total RNA Sample Preparation kits and all the ribosomal RNA was depleted using Ribo-Zero Gold. Sequencing was performed on the Illumina Hi-Seq 2500 using a 100bp paired-end protocol at the Flinders University Genomics Facility and South Australian Health & Medical Research Institute (SAHMRI).

### Data processing and statistical analysis

All sequencing data was firstly assessed for quality using FastQC and ngsReports(*28, 29*) to identify any failed and low-coverage samples. Raw paired-end, stranded FASTQ files were then trimmed for sequence adapters using AdapterRemoval(*30*) and aligned to the 1000 genomes project GRCh37 (hs37d5) human reference genome using STAR(*31*) and gene counts were identified on raw alignments using featureCounts(*32*). Analysis was limited to gene counts greater than 2 counts per million (CPM) in at least 55 samples (i.e. the size of the smallest sample group).

Differential gene expression (DGE) was carried out using edgeR(*33*) within the R/Bioconductor statistical framework, relying on sample-weight transformation contained in the *voom* functions(*34*). Linear models were then fitted with *limma(35)* along with empirical Bayes moderation.

### Co-expression network and gene set enrichment

Weighted gene correlation network analysis (WGCNA) was carried out using R package WGCNA(*36*) using two sets of early gestation RNA-seq samples. Low counts were removed similar to DGE, variance stabilisation and batch effects removed using *DESeq2(37)* and low variability genes removed (i.e. genes in the lowest quartile of standard deviation). Signed network and adjacency matrix were constructed using a soft power threshold of 14 and modules with a minimum size of 50 generated and merged at distance cut-off of 0.25. Network graphs were constructed and visualised using Gephi (https://gephi.org/).

Module and differentially expressed genes were tested for enrichment within Gene Ontology and Molecular Signature Database (MSigDB v6.2) gene sets(*38*) using the WGCNA R package *AnRichment* (https://horvath.genetics.ucla.edu/html/CoexpressionNetwork/GeneAnnotation/) and *clusterProfiler(39)*. Additional single sample enrichment was carried out using the package *singscore(12)* using cell-line and tumour cell-line Epithelial-Mesenchymal Transition (EMT) gene sets derived from (*13*).

## Data Availability

Full code and workflow available (https://github.com/jimmybgammyknee/earlyPlacentaRnaSeqProfile). All sequencing data is available for download at NCBI Gene Expression Omnibus (GEO) under Accession number: PRJNA633801.

## Acknowledgments

We thank the women who kindly donated their placenta tissue and the clinical staff who cared for them. Additional thanks goes to other researchers that gave useful information, including John Schjenken, Sam Buckberry and Dan Kortschak.

## Author contributions

JB, TBM and CTR devised the project. DaM, TJB and DyM conducted sampling and wet laboratory work. JB conducted the analyses with input from MDS, AC, NH and SP. JB and CTR interpreted the data. JB wrote the manuscript with intellectual input from MDS, TBM and CTR. All authors approved the final manuscript.

